# Comprehensive Evaluation of Associations between Lifestyle Factors and Multiple Epigenetic Aging Indicators in the Japanese Population: A cross-sectional study

**DOI:** 10.64898/2026.02.07.26345813

**Authors:** Tatsuma Shoji, Genki Yoshikawa, Sawako Hibino, Hidekazu Yamada, Ryo Nakaki

## Abstract

**Background:** Epigenetic clocks based on DNA methylation (DNAm) provide quantitative indicators of biological aging. However, the extent to which diverse lifestyle factors influence DNAm-based aging measures remains unclear, especially in Japanese populations. We aimed to evaluate the associations between 52 lifestyle-related factors and multiple epigenetic aging indicators, including six DNAm ages (Horvath, Hannum, PhenoAge, GrimAge, GrimAge v2, and PCPhenoAge specific to Japanese Population), the Dunedin PACE, and six corresponding age acceleration indices. We recruited 287 Japanese adults between January and December 2024 and evaluated the association of these aging indices with their responses on lifestyle questionnaires using multivariable linear regression models. We entered items either individually or simultaneously while adjusting for major confounding factors.

**Results:** DNAm ages showed strong intercorrelations, whereas age acceleration indices demonstrated weaker correlations. Several lifestyle factors such as late-night eating, processed food intake, smoking-related behaviors, high-intensity interval training, and thermal relaxation habits exhibited strong and clock-specific associations with aging indices. In simultaneous models incorporating all 52 factors, only a limited subset, with factors such as smoking exposure, high-intensity exercise, and sauna or stone spa use, were associated with aging indices. Each DNAm aging indicator demonstrated a distinct pattern of association with lifestyle exposure, indicating that epigenetic aging indices capture different physiological processes.

**Conclusions:** These findings may improve our understanding of lifestyle–epigenetic interactions and provide evidence supporting the use of DNAm-based biological age as a tool for personalized healthcare in Japan.

## Background

Lifestyle behaviors contribute to the onset and progression of various chronic diseases, including cardiovascular diseases, type 2 diabetes, obesity, chronic obstructive pulmonary disease, and malignant tumors [1]. Most of these conditions depend on modifiable behavioral factors such as unhealthy dietary patterns, insufficient physical activity, cigarette smoking, and excessive alcohol consumption [2]. In addition to the rapid aging of the Japanese population, rising medical costs, driven by lifestyle-related diseases and the stagnation of healthy life expectancy have become major societal issues [3]. Consequently, improving individual health literacy and promoting preventive medicine have become important issues [4]. Moreover, there is a rising demand for biomarkers that link lifestyle behaviors to health risks. Epigenetic clocks, which estimate biological age, distinct from chronological age, based on DNA methylation patterns, have attracted growing attention [5]. Representative models include the pan-tissue clock proposed by Horvath [6], the blood-specific model developed by Hannum [7], PhenoAge reflecting health span [8], GrimAge with high predictive accuracy for mortality risk [9], and the Dunedin PACE, which estimates the pace of aging [10]. Furthermore, a Japan-specific model has been developed using a transfer learning approach [11]. In this study, we refer to the PC-PhenoAge for Japanese Population as the J-PCPhenoAge. These models and the estimated biological ages reflect lifestyle behaviors and may be applied as intervention metrics to promote behavioral modifications [12].

Although epigenetic clocks are widely used to assess biological aging and disease risk, comprehensive investigations examining their relationship with lifestyle behaviors remain limited [13]. Previous studies have focused on individual clocks, and few have compared multiple clocks within the same population while accounting for the differences in their characteristics [14]. Moreover, most existing evidence has been generated from Western populations, and there is a lack of data from East Asian groups, particularly Japanese individuals, whose genetic backgrounds, dietary patterns, occupational structures, and social security systems differ [5]. Particularly, evidence regarding newer indicators such as GrimAge v2 and Dunedin PACE is scarce [15]. Although J-PCPhenoAge [11], has been proposed, few studies have compared it to established clocks. Therefore, there is a need for research that compares multiple epigenetic clocks and clarifies how these clocks are associated with lifestyle behaviors in Japanese individuals.

The aim of this study was to evaluate the association between multiple epigenetic aging indicators and lifestyle behaviors in Japanese adults. We sought to elucidate how each aging indicator relates to lifestyle behaviors among Japanese individuals, and to identify which indicators capture behavioral characteristics.

## Methods

### 1) Study Design and Participants

This observational cross-sectional study involved adults who visited Y’s Science Clinic Hiroo (Minato-ku, Tokyo, Japan) between January and December 2024. Clinic attendees were recruited to participate in this study. Written informed consent was obtained from all participants. A total of 287 individuals were initially enrolled, and all were included in the final analysis (N = 287). The study protocol was approved by the Ethics Committee of the Asia–Oceania Association for Anti-Aging Promotion (approval number: AOAAPA22-001).

We evaluated the associations of six epigenetic clocks (Horvath, Hannum, PhenoAge, GrimAge, GrimAge v2, and J-PCPhenoAge), an aging pace indicator (Dunedin PACE), and corresponding epigenetic age acceleration (AgeAccel) indices within the same population with various lifestyle factors, including dietary habits, and alcohol consumption, physical activity, smoking, sleep, stress, and social factors.

### 2) Lifestyle Assessment

Information on lifestyle factors was obtained using a self-reported questionnaire (Additional file 1). The questionnaire was designed to comprehensively capture lifestyle behaviors among Japanese adults and categorized into five domains: Food (dietary habits and alcohol consumption), Sleep, Exercise, Smoking, and Others. Each item was selected to cover behavioral factors previously reported to be associated with chronic diseases and epigenetic aging, thereby ensuring the relevance of the questionnaire to aging-related health research [16–18].

The Food category included items related to the basic temporal structure of meals, such as “Breakfast Habit,” “Lunch Habit,” “Dinner Habit,” ”Snack Habit,” and “Late night Snack Habit,” as well as items assessing the regulation or restriction of energy intake, such as “Caloric Intake Management,” “Caloric Restriction Frequency,” and “Carbohydrate Restriction.” Additionally, the questionnaire covered the consumption of specific foods and beverages, including “Daily Vegetable Intake,” “Daily Fruit Intake,” “Weekly Nut Intake,” “Weekly Yogurt Kefir Consumption,” “Weekly Ultra processed Food Consumption,” “Weekly Omega 3 Fatty Acid Food Intake,” “Weekly Poultry Consumption,” “Weekly Red Meat Consumption,” “Weekly Refined Grain Intake,” “Weekly Whole Grain Intake,” “Daily Lactic Acid Beverage Intake,” “Daily Coffee Intake,” “Daily Tea Intake,” “Daily Soft Drink Intake,” and “Daily Water Intake,”. Furthermore, “Use of Artificial Sweeteners in Food and Drink,” “Weekly Alcohol Consumption Days,” “Daily Alcohol Intake,” and “Supplement Intake,” were included to assess the use of food additives, alcohol, and supplements. The Sleep category included “Quality of Sleep and Rest,” “Average Bedtime in the Past Year,” “Average Sleep Duration,” “Difference in Sleep Duration Between Weekdays and Weekends,” “Sleep Quality in the Past Month,” “Relaxation 30 Minutes Before Bedtime,” and “Caffeine Consumption Within 6 h Before Bedtime.” The Exercise category included “Weekly Stretching Exercise Duration”, “Weekly Low intensity Resistance Exercise Duration”, “Weekly Moderate intensity Resistance Exercise Duration,” “Weekly Low intensity Aerobic Exercise Duration,” “Weekly Moderate intensity Aerobic Exercise Duration,” “Weekly High intensity Interval Training Duration,” “Weekly Physical Labor Duration,” “Daily Sitting Time,” “Weekly Sauna Bathing Duration,” and “Weekly Stone Spa Bathing Duration.” The Smoking category included “Cigarette Smoking,” “Electronic Cigarette Smoking,” “Past Smoking Habit,” and “Past Frequent Secondhand Smoke Exposure,” comprehensively assessing current smoking, e-cigarette use, past smoking behavior, and exposure to secondhand smoke. The Others category included “Time Spent in Nature,” “Daily Direct Sunlight Exposure Duration,” “Pet Ownership,” and “Frequency of Dental Checkups.” These items represent factors that reflect the social and environmental context of daily life in Japan. Additionally, the questionnaire included items on sex, chronological age, educational attainment, and household income. These items reflect socioeconomic background and physical characteristics that may influence both lifestyle behaviors and biological aging; therefore, they were considered confounders [19].

The response data are summarized in Additional file 2. All responses were binarized according to the following criteria: For food-related items, such as “Caloric Intake Management,” “Caloric Restriction Frequency,” and “Carbohydrate Restriction,” a value of 1 was assigned for engagement in these practices. For all other food items, a value of 1 was assigned for consumption of the respective food or beverage. For sleep-related items, “Average Bedtime in the Past Year” was coded as 1 for responses indicating a bedtime ≥ 00:00. “Average Sleep Duration” was coded as 1 for a duration ≥6 h. Sleep Quality in the Past Month was coded as 1 for responses at or above “High.” All the remaining sleep-related items were coded as 1 when the participant reported experiencing the respective condition or behavior. For exercise-related items, Daily Sitting Time was assigned a value of 1 for sitting durations ≥8 h, whereas the other exercise items were assigned a value of 1 when the respective activity was reported. For the four smoking-related items, a value of 1 was assigned when the participant reported a corresponding behavior. Among the “Others” items, “Frequency of Dental Checkups” was coded as 1 when participants reported undergoing dental examinations at least once per year. All the remaining items in this category were coded as 1 when the corresponding behavior or condition was reported (Additional file 3). 3) Blood Sample Collection and DNA Methylation Measurement

Blood sample collection and DNA methylation analyses were performed as previously described [20]. Briefly, blood samples were collected from Y’s Science Clinic Hiroo using EDTA tubes, stored at 4 °C, and processed promptly. Genomic DNA was extracted using the Maxwell RSC Blood DNA Kit. DNA methylation was measured using Illumina Infinium arrays (MethylationEPIC v2.0 and Methylation Screening Array), which were scanned on an Illumina iScan system at the Rhelixa. The raw IDAT files were processed in R (v4.4.2) using the SeSAMe pipeline (v1.24.0) [21, 22]. Beta values were computed as the ratio of methylated to total signal [23], and established epigenetic clocks (Horvath, Hannum, PhenoAge, GrimAge, GrimAge v2, DunedinPACE, and J-PCPhenoAge) were calculated from the processed data.

### 4) Calculation of Epigenetic Clocks

Blood sample collection and DNA methylation analyses were performed as previously described [20]. Briefly, blood samples were collected from Y’s Science Clinic Hiroo using EDTA tubes, stored at 4 °C, and processed promptly. Genomic DNA was extracted using the Maxwell RSC Blood DNA Kit. DNA methylation was measured using Illumina Infinium arrays (MethylationEPIC v2.0 and Methylation Screening Array), which were scanned on an Illumina iScan system at the Rhelixa. The raw IDAT files were processed in R (v4.4.2) using the SeSAMe pipeline (v1.24.0) [21, 22]. Beta values were computed as the ratio of methylated to total signal [23], and established epigenetic clocks (Horvath, Hannum, PhenoAge, GrimAge, GrimAge v2, DunedinPACE, and J-PCPhenoAge) were calculated from the processed data.

### 5) Statistical Analysis

The 52 lifestyle-related factors used in this study were defined as categorical variables All explanatory variables were binary (0 or 1) and descriptive statistics are presented as percentages. In the analyses, the DNA methylation age (DNAmAge) standardized by z-score transformation or AgeAccel of each epigenetic clock, computed using scikit-learn (version 1.7.1) in Python (version 3.12.3), was used as the dependent variable.

We specified two types of models: (i) each lifestyle factor was individually entered into a multivariate linear regression model, with major confounders included as covariates (sex, chronological age [in the standardized DNAmAge model only], educational attainment, and household income); and (ii) all lifestyle variables and confounders were simultaneously entered into multivariate linear regression models. The models were fitted to males and females separately. The primary evaluation metrics were the estimated regression coefficients, 95% confidence intervals, p-values, and effect sizes, calculated as the regression coefficient divided by the standard deviation (SD) of the model residuals (a measure analogous to Cohen’s d). Missing data were handled using multiple imputations [24–26] using the mice package (version 3.17.0) in R (version 4.3.1), generating 50 imputed datasets with five iterations. Linear regression models were fitted to each imputed dataset using the stats package (version 4.3.1) and estimates were pooled according to Rubin’s rules. These metrics were used to assess the extent to which each lifestyle factor independently influenced the epigenetic indicators. In this study, lifestyle factors with an effect size > 0.5 and those that showed a balanced response, defined as cases in which one category accounted for < 95% of the responses after binarization, were considered to be associated. Since this was an exploratory study, no adjustments for multiple comparisons were applied. Heatmaps were created using seaborn (version 0.13.2) in Python (version 3.12.3).

## Results

### 1) Participant Characteristics

A total of 287 individuals participated in this study. The age distribution was as follows: 10–19 years, 1 participant (0.35%); 20–29 years, 29 participants (10.10%); 30–39 years, 59 participants (20.56%); 40–49 years, 83 participants (28.92%); 50–59 years, 63 participants (21.95%); 60–69 years, 35 participants (12.20%); 70–79 years, 16 participants (5.57%); and 80–89 years, 1 participant (0.35%). The sample comprised 158 males (55.05%) and 129 females (44.95%). Questionnaire response rates across all categories were high: 91.61% for Food; 98.26% for Sleep, Exercise, and Smoking; and 95.38% for Others (Table 1).

**Table 1.**
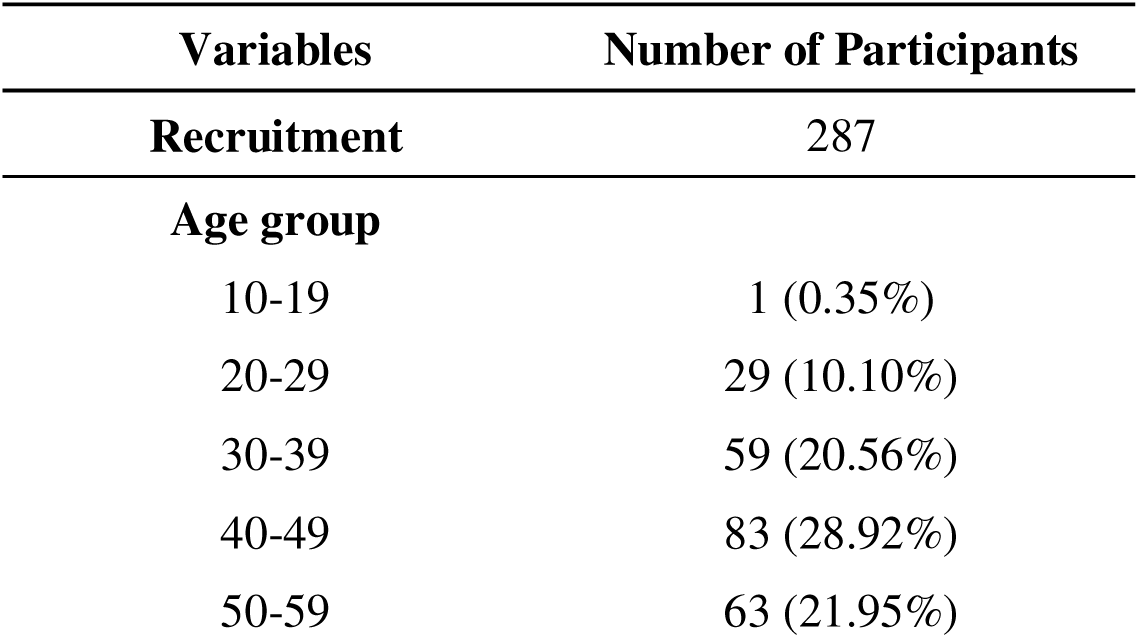
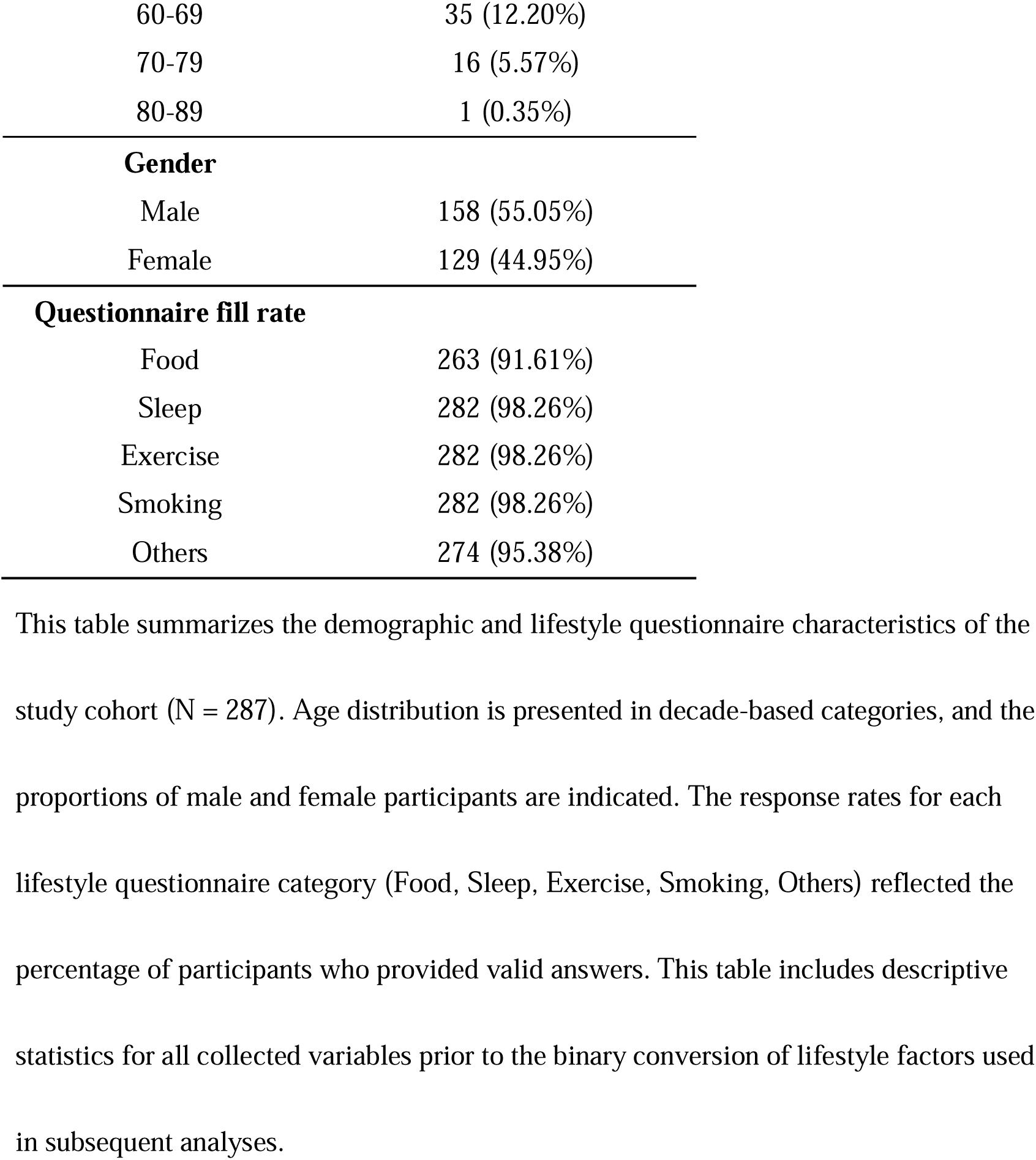
Participant characteristics.

Within the Food category, lunch habit (96.45%), dinner habit (99.29%), vegetable intake (89.72%), refined grain intake (92.20%), and water intake (94.33%) were highly prevalent, whereas the use of artificial sweeteners (7.80%) and high-intensity exercise (13.12%) were infrequent. In the Sleep category, awareness of good sleep and rest (62.41%), adequate average sleep duration (60.64%), and relaxation 30 min before bedtime (60.99%) were observed. In the Exercise category, stretching (60.64%) and low-intensity aerobic exercise (57.45%) were common, whereas sauna treatment (18.44%) and stone spa bathing (3.55%) were less frequent. In the Smoking category, current cigarette smoking, electronic cigarette use, previous smoking history, and frequent previous secondhand smoke exposure were reported by 7.80%, 11.70%, 45.04%, and 29.79%, respectively. In the Others category, the proportions of participants who reported spending time in nature (83.33%) and being exposed to sunlight (69.68%) were high, whereas that of those who reported owning pets was relatively low (33.59%) (Table 2).

**Table 2.** Distribution of responses to the 52 lifestyle-related questionnaire items across five categories.

| **Lifestyle-Related Factors (Food)** | **N** | **Percentage** |
| --- | --- | --- |
| **Breakfast Habit** | 241 | 85.46% |
| **Lunch Habit** | 272 | 96.45% |
| **Dinner Habit** | 280 | 99.29% |
| **Snack Habit** | 227 | 80.50% |
| **Late night Snack Habit** | 114 | 40.43% |
| **Caloric Intake Management** | 125 | 44.33% |
| **Caloric Restriction Frequency** | 42 | 14.63% |
| **Carbohydrate Restriction** | 39 | 35.45% |
| **Daily Vegetable Intake** | 253 | 89.72% |
| **Daily Fruit Intake** | 144 | 51.06% |
| **Weekly Nut Intake** | 96 | 34.04% |
| **Weekly Yogurt Kefir Consumption** | 153 | 54.26% |
| **Weekly Ultra processed Food Consumption** | 169 | 59.93% |
| **Weekly Omega 3 Fatty Acid Food Intake** | 221 | 78.37% |
| **Weekly Poultry Consumption** | 251 | 89.01% |
| **Weekly Red Meat Consumption** | 256 | 90.78% |
| **Weekly Refined Grain Intake** | 260 | 92.20% |
| **Weekly Whole Grain Intake** | 140 | 49.65% |
| **Daily Lactic Acid Beverage Intake** | 128 | 45.39% |
| **Daily Coffee Intake** | 230 | 81.56% |
| **Daily Tea Intake** | 90 | 31.91% |
| **Daily Soft Drink Intake** | 90 | 31.91% |
| **Daily Water Intake** | 266 | 94.33% |
| **Use of Artificial Sweeteners in Food and Drink** | 22 | 7.80% |
| **Weekly Alcohol Consumption Days** | 179 | 63.48% |
| **Daily Alcohol Intake** | 179 | 63.48% |
| **Supplement Intake** | 160 | 56.74% |

| **Lifestyle-Related Factors (Sleep)** | **N** | **Percentage** |
| --- | --- | --- |
| **Quality of Sleep and Rest** | 176 | 62.41% |
| **Average Bedtime in the Past Year** | 144 | 51.06% |
| **Average Sleep Duration** | 171 | 60.64% |
| **Difference in Sleep Duration Between Weekdays and Weekends** | 168 | 59.57% |
| **Sleep Quality in the Past Month** | 112 | 39.72% |
| **Relaxation 30 Minutes Before Bedtime** | 172 | 60.99% |
| **Caffeine Consumption Within 6 Hours Before Bedtime** | 158 | 56.03% |

| **Lifestyle-Related Factors (Exercise)** | **N** | **Percentage** |
| --- | --- | --- |
| **Weekly Stretching Exercise Duration** | 171 | 60.64% |
| **Weekly Low intensity Resistance Exercise Duration** | 137 | 48.58% |
| **Weekly Moderate intensity Resistance Exercise Duration** | 100 | 35.46% |
| **Weekly Low intensity Aerobic Exercise Duration** | 162 | 57.45% |
| **Weekly Moderate intensity Aerobic Exercise Duration** | 101 | 35.82% |
| **Weekly High intensity Interval Training Duration** | 37 | 13.12% |
| **Weekly Physical Labor Duration** | 51 | 18.09% |
| **Daily Sitting Time** | 105 | 37.23% |
| **Weekly Sauna Bathing Duration** | 52 | 18.44% |
| **Weekly Stone Spa Bathing Duration** | 10 | 3.55% |
| **Lifestyle-Related Factors (Smoking)** | **N** | **Percentage** |
| **Cigarette Smoking** | 22 | 7.80% |
| **Electronic Cigarette Smoking** | 33 | 11.70% |
| **Past Smoking Habit** | 127 | 45.04% |
| **Past Frequent Secondhand Smoke Exposure** | 84 | 29.79% |
| **Lifestyle-Related Factors (Others)** | **N** | **Percentage** |
| **Time Spent in Nature** | 235 | 83.33% |
| **Daily Direct Sunlight Exposure Duration** | 193 | 69.68% |
| **Pet Ownership** | 87 | 33.59% |
| **Frequency of Dental Checkups** | 211 | 76.17% |

For each item, the table reports the number of participants who met the criteria for the
binary variable (N) and the corresponding percentage relative to the total number of
respondents for that item. Higher percentages indicate more prevalent lifestyle habits
within the study population. The Food category covers dietary patterns (meal timing,
nutrient intake, specific food groups, beverages, additives, and alcohol/supplement use).
The Sleep category includes sleep quality, sleep duration, bedtime patterns, and pre-sleep
behaviors. The Exercise category comprises stretching, resistance/aerobic exercise at
multiple intensities, physical labor, sitting time, sauna bathing, and stone spa bathing. The
Smoking category covers current smoking, electronic cigarette use, past smoking, and

DNAmAge, expressed as mean and SD, were 52.29 ± 12.00, 18.09 ± 11.52, 42.56 ± 14.04, 48.66 ± 11.08, 53.81 ± 10.92, and 42.26 ± 12.93 years for Horvath, Hannum, PhenoAge, GrimAge, GrimAge v2, and J-PCPhenoAge, respectively. They all showed strong correlations with chronological age (r = 0.911–0.958). The DunedinPACE value (mean ± SD) was 1.02 ± 0.09, with a moderate correlation with chronological age (r = 0.456). For AgeAccel indices, the means and SDs were 0.00 ± 3.89, 0.00 ± 3.09, 0.00 ± 5.80, 0.00 ± 3.20, 0.00 ± 3.70, and 0.00 ± 4.29 for Horvath, Hannum, PhenoAge, GrimAge, GrimAge v2, and J-PCPhenoAge, respectively (Table 3). Based on the classification of individuals with biological ages above the chronological age (Table 4), 89.20% of the participants exceeded their chronological age according to the Horvath Clock, 60.63% according to GrimAge, and 11.15% according to J-PCPhenoAge.

**Table 3.** Descriptive statistics for the 13 epigenetic aging indicators.

| **Biological Clocks** | **Mean** | **SD** | **Minimum** | **Maximum** | **Correlation with**<br>**Chronological Age** |
| --- | --- | --- | --- | --- | --- |
| Horvath | 52.29 | 12.00 | 21.22 | 84.13 | 0.944 |
| Hunnum | 18.09 | 11.52 | -9.84 | 48.83 | 0.958 |
| PhenoAge | 42.56 | 14.04 | 5.55 | 82.61 | 0.911 |
| GrimAge | 48.66 | 11.08 | 24.59 | 79.09 | 0.95 |
| GrimAge.v2 | 53.81 | 10.92 | 29.55 | 84.09 | 0.936 |
| J-PCPhenoAge | 42.26 | 12.93 | 8.21 | 79.13 | 0.943 |
| DunedinPACE | 1.02 | 0.09 | 0.78 | 1.36 | 0.456 |
| **Accelerated aging**<br>**(AA)** |  |  |  |  |  |
| Horvath AA | 0.00 | 3.89 | -13.93 | 12.79 | - |
| Hunnum AA | 0.00 | 3.09 | -9.00 | 10.83 | - |
| PhenoAge AA | 0.00 | 5.80 | -28.55 | 20.51 | - |
| GrimAge AA | 0.00 | 3.20 | -6.80 | 12.26 | - |
| GrimAge.v2 AA | 0.00 | 3.70 | -8.56 | 16.20 | - |
| J-PCPhenoAge AA | 0.00 | 4.29 | -10.45 | 14.99 | - |

This table summarizes the distribution of each epigenetic aging indicator in the study population. For DNAmAge measures (Horvath, Hannum, PhenoAge, GrimAge, GrimAge v2, J-PCPhenoAge, and DunedinPACE), the table reports the mean and standard deviations of the estimated epigenetic age (or aging pace for DunedinPACE). Age acceleration (AgeAccel) indicators were computed as residuals from the linear regression of DNAmAge on chronological age, and their distribution characteristics (mean and standard deviation) are shown separately. These descriptive statistics provide an overview of the inter-individual variability in both absolute epigenetic age levels and age-adjusted biological aging rates across 287 participants.

**Table 4.**
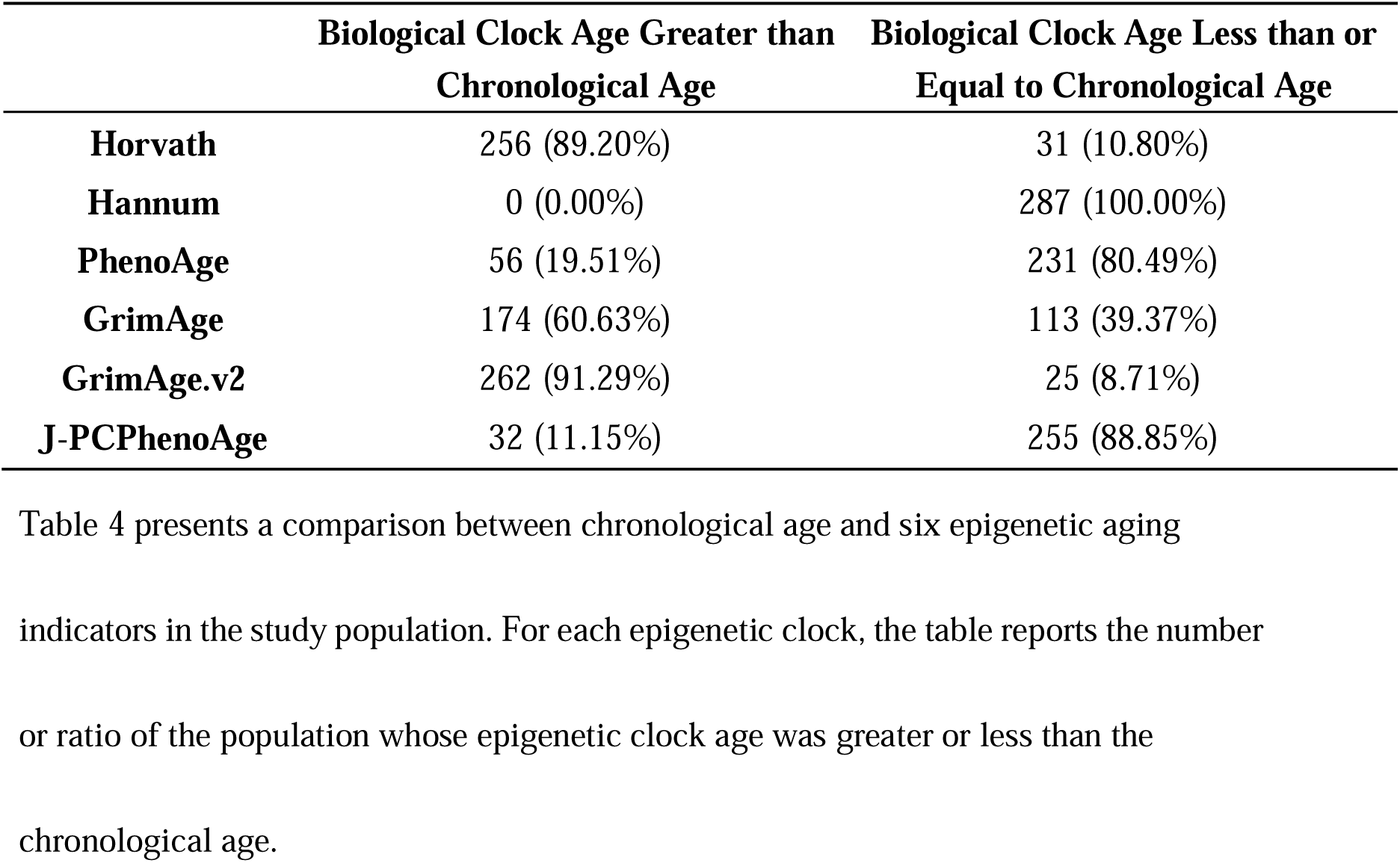
Comparison Between Chronological Age and Epigenetic Age.

### 3) Associations between Individual Lifestyle Factors and Epigenetic Aging

As shown in Figure 1A, consistently strong positive correlations were observed among DNAmAge measures. The correlation coefficients between Horvath, Hannum, PhenoAge, GrimAge, GrimAge v2, and J-PCPhenoAge ranged from r = 0.91 to 0.99, suggesting that multiple clocks capture common age-related methylation patterns. The strongest correlation was observed between GrimAge and GrimAge v2 (r = 0.99), followed by Hannum and GrimAge (r = 0.95), and J-PCPhenoAge and Hannum (r = 0.95). Contrarily, the correlations among AgeAccel measures (Figure 1B) were weaker and more heterogeneous than those among DNAmAge measures. Moderate associations were observed for pairs such as Horvath AA and Hannum AA (r = 0.52) and PhenoAge AA and J-PCPhenoAge AA (r = 0.62), whereas the correlation between Horvath AA and GrimAge AA was low (r = 0.21). GrimAge AA and GrimAge v2 AA exhibited a very strong correlation (r = 0.96).

**Figure 1.**
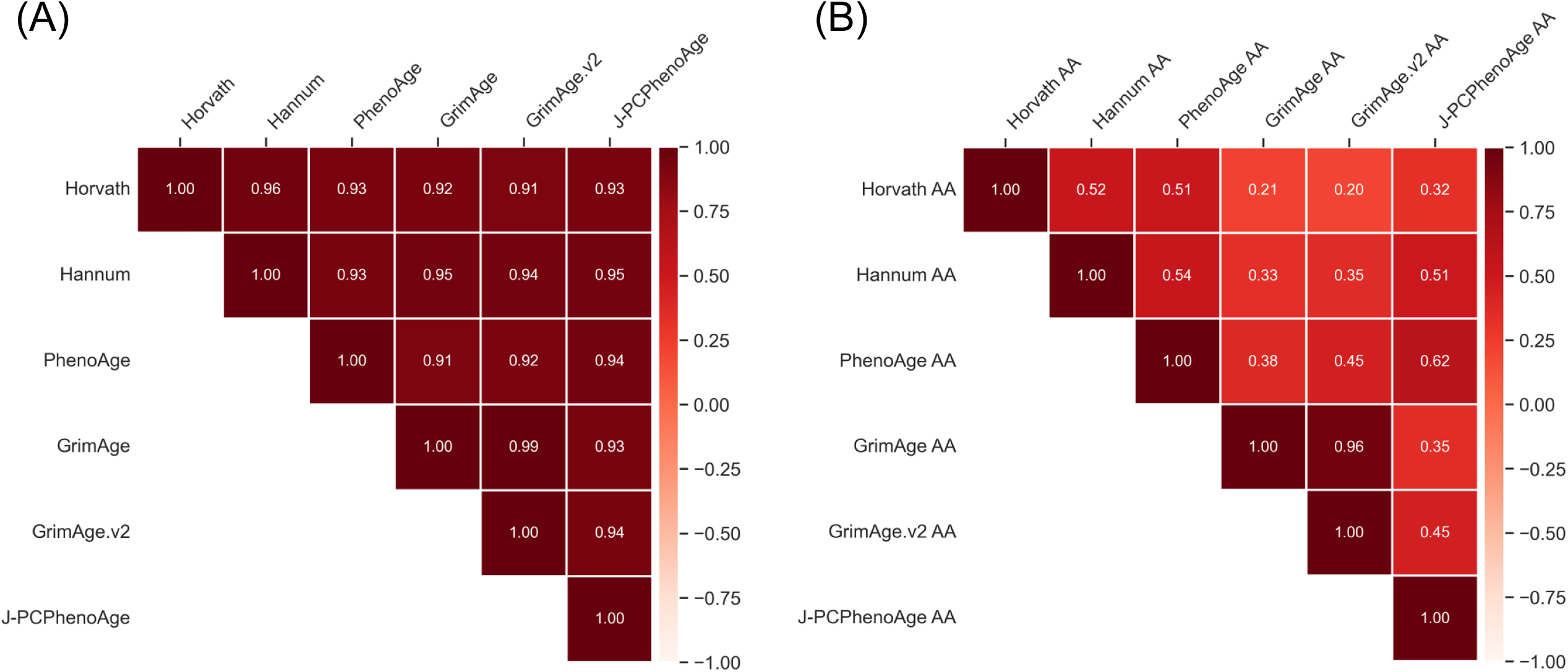
Pearson correlation matrices among epigenetic aging indicators. **(a** Correlations among seven DNAmAge measures (Horvath, Hannum, PhenoAge, GrimAge, GrimAge v2, J-PCPhenoAge). Strong positive correlations were observed across most DNAmAge indicators, reflecting shared methylation-based aging signatures. **(b)** Correlations among six age acceleration (AgeAccel) measures derived from the corresponding DNAmAge indicators. AgeAccel measures showed lower and more heterogeneous correlations compared with DNAmAge, consistent with their residualized nature after adjusting for chronological age. Color gradients represent correlation coefficients, with darker shades indicating stronger positive correlations.

### 3) Associations between Individual Lifestyle Factors and Epigenetic Aging

There were strong associations between DNAmAge and multiple lifestyle factors, with clear sex-specific patterns. In males, the associations were observed between AgeAaccel and behaviors such as smoking, irregular meal timing, use of artificial sweeteners, alcohol consumption, poultry intake, pet ownership, and high water intake. Meanwhile, vegetable intake and stone spa bathing showed consistent age-decelerating effects. In females, electronic cigarette use, combustible cigarette smoking, frequent caloric restriction, carbohydrate restriction, snacking habits, and certain meal-timing behaviors, were associated with DNAm AgeAccel; breakfast consumption and high-intensity interval training (HIIT) demonstrated protective effects (Figures 2 and 3, Additional file 4 and 5).

**Figure 2.**
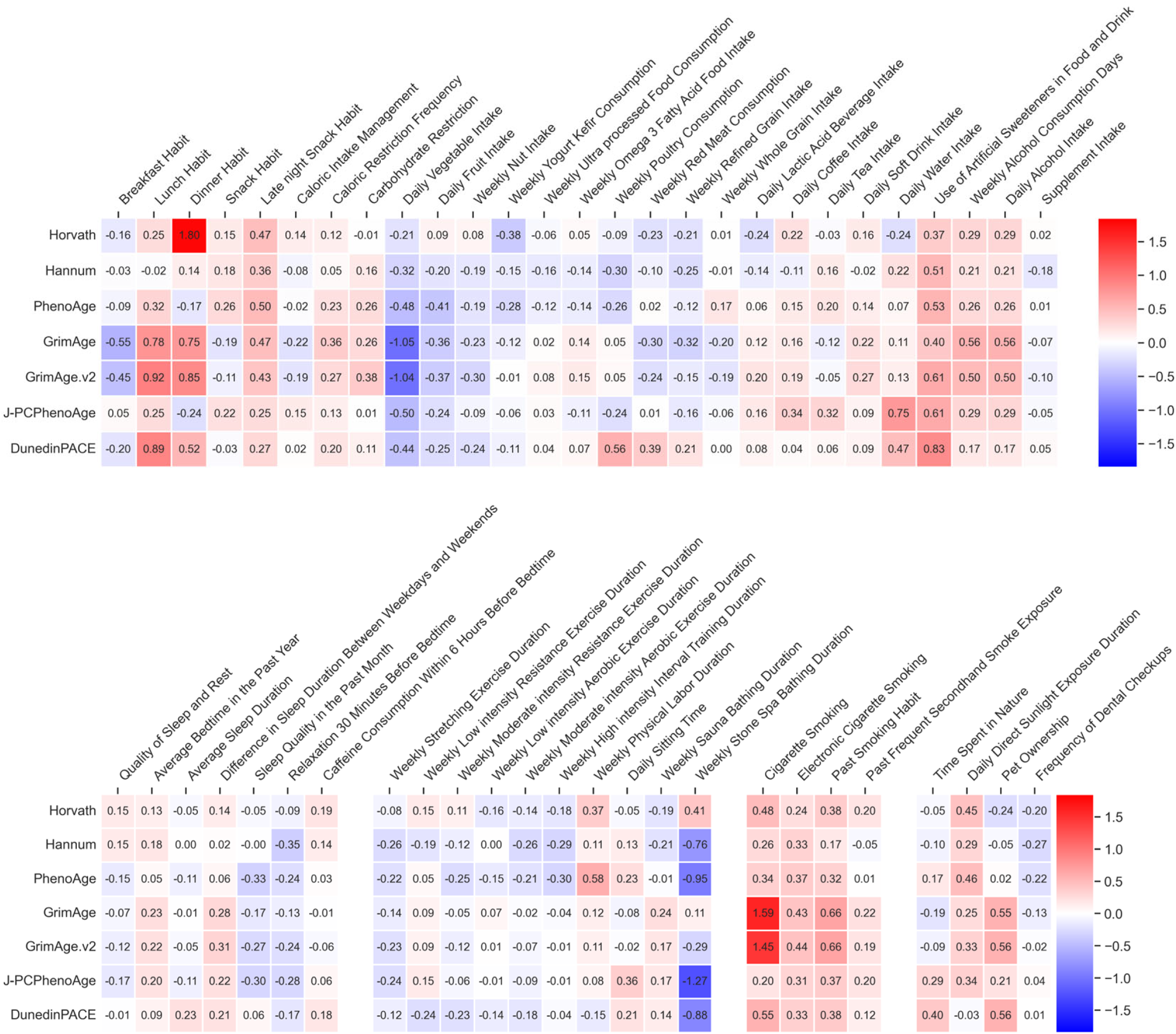
Heatmap of effect sizes for associations between seven DNAmAge indicators and 52 lifestyle-related factors in males. Each cell represents the standardized effect size derived from multivariable linear regression models in which one lifestyle factor was entered as the explanatory variable and a DNAmAge indicator (Horvath, Hannum, PhenoAge, GrimAge, GrimAge v2, J-PCPhenoAge, or DunedinPACE) was used as the dependent variable. Only the data for males are shown in this figure.

**Figure 3.**
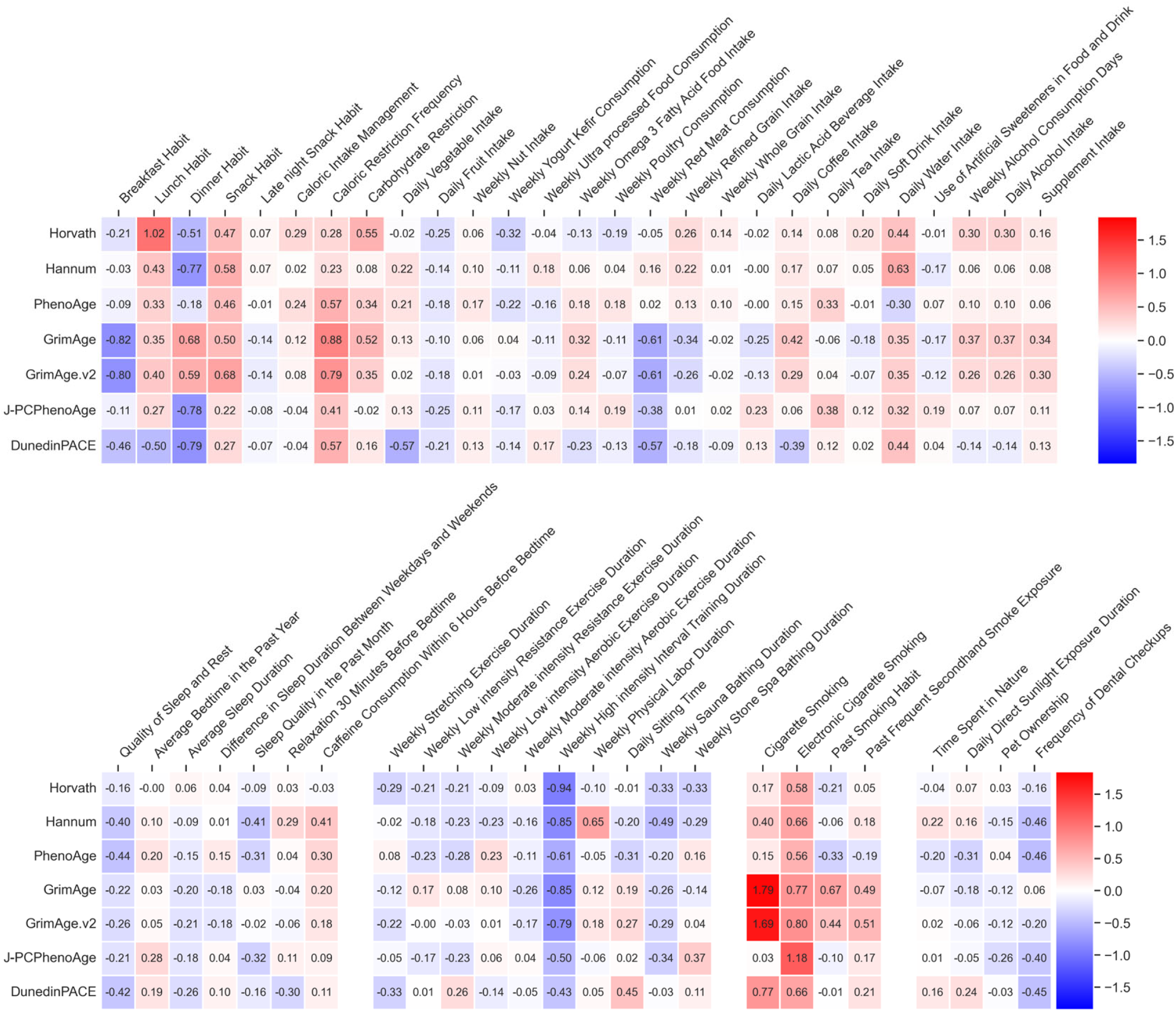
Heatmap of effect sizes for associations between seven DNAmAge indicators and 52 lifestyle-related factors in females. Each cell represents the standardized effect size derived from multivariable linear regression models in which one lifestyle factor was entered as the explanatory variable and a DNAmAge indicator (Horvath, Hannum, PhenoAge, GrimAge, GrimAge v2, J-PCPhenoAge, or DunedinPACE) was used as the dependent variable. Only the data for females are shown in this figure.

AgeAccel was strongly associated with smoking behavior, alcohol consumption, eating patterns, snacking, caloric restriction, and carbohydrate restriction in both males and females. In males, cigarette smoking showed the strongest association with epigenetic aging, followed by artificial sweetener use, physical labor, and water intake. Conversely, vegetable intake and stone spa bathing were negatively associated with epigenetic aging in both sexes; stone spa bathing showed the strongest association in males. Among females, electronic cigarette use had the strongest association across multiple clocks, whereas HIIT and regular breakfast consumption showed negative associations, highlighting a clearer pattern of responsiveness to smoking, dietary behaviors, and physical activity. Overall, AgeAccel exhibited stronger associations with lifestyle factors than DNAmAge, capturing distinct behavioral signatures related to smoking, dietary patterns, physical activity, and thermal relaxation practices (Figures 4 and 5, Additional file 6 and 7).

**Figure 4.**
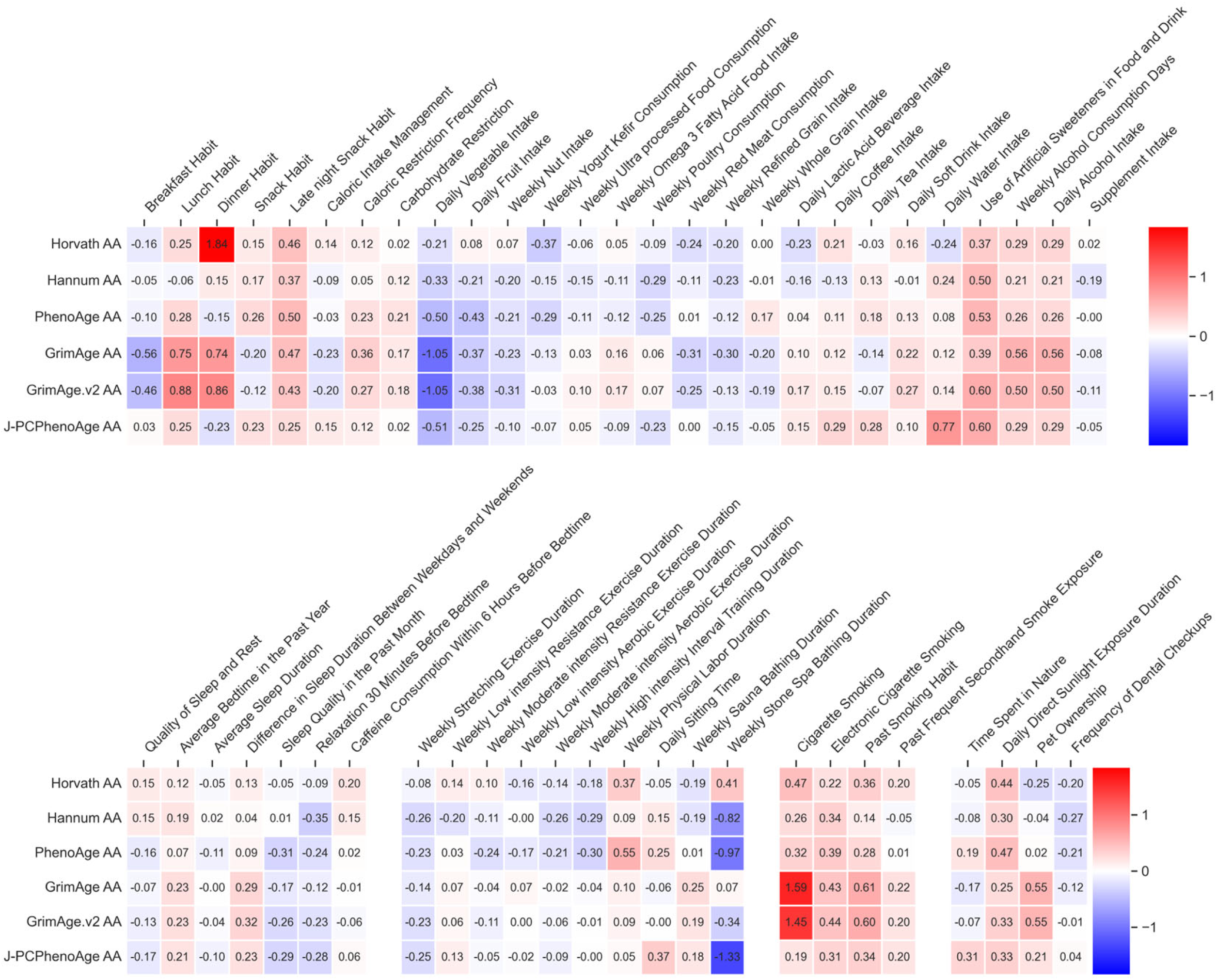
Heatmap of effect sizes for associations between six AgeAccel indicators and 52 lifestyle-related factors in males. Each cell represents the standardized effect size derived from multivariable linear regression models, in which one lifestyle factor was entered as the explanatory variable and an AgeAccel indicator (Horvath AA, Hannum AA, PhenoAge AA, GrimAge AA, GrimAge v2 AA, or J-PCPhenoAge AA) was used as the dependent variable. Only the data for males are shown in this figure.

**Figure 5.**
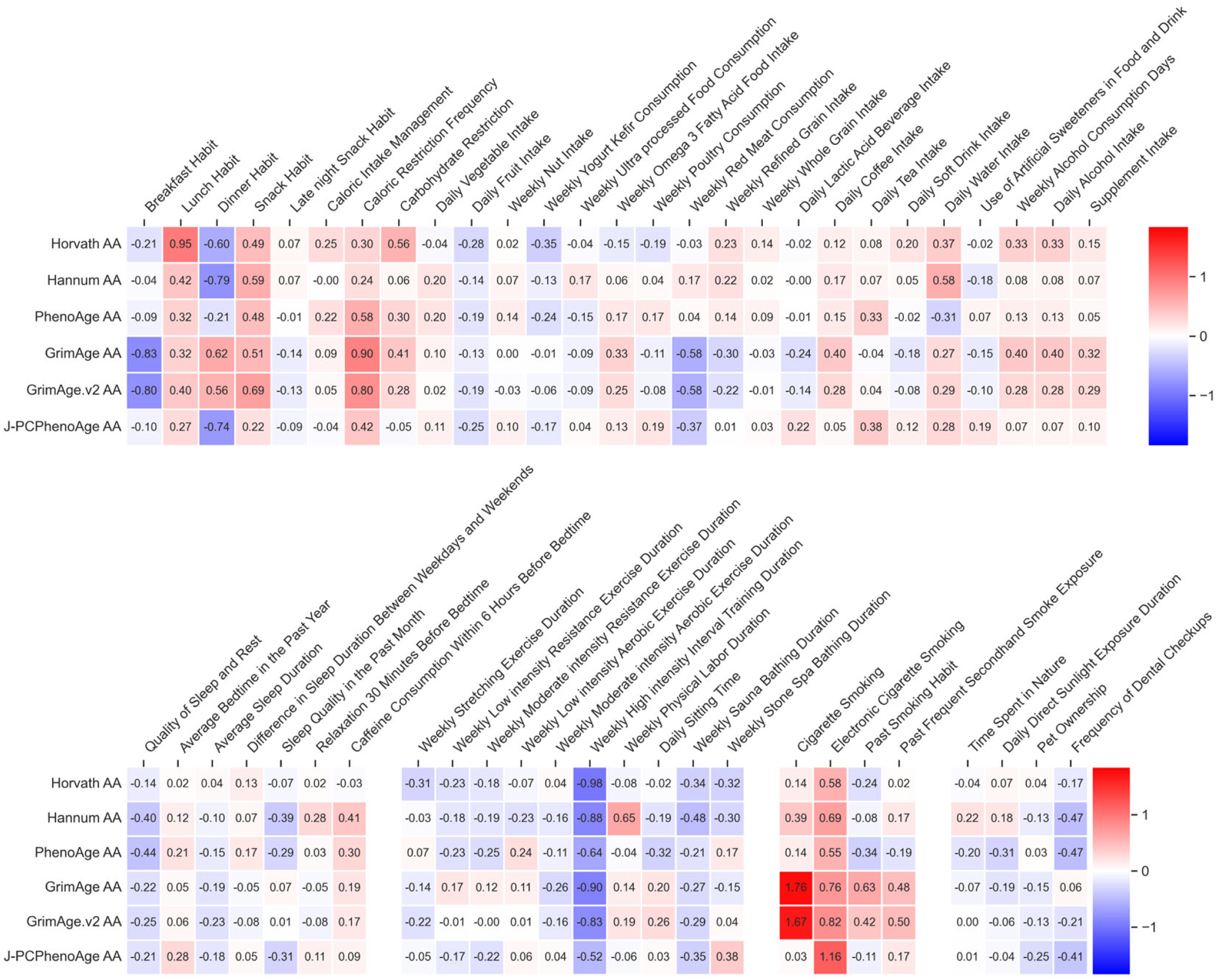
Heatmap of effect sizes for associations between six AgeAccel indicators and 52 lifestyle-related factors in females. Each cell represents the standardized effect size derived from multivariable linear regression models, in which one lifestyle factor was entered as the explanatory variable and an AgeAccel indicator (Horvath AA, Hannum AA, PhenoAge AA, GrimAge AA, GrimAge v2 AA, or J-PCPhenoAge AA) was used as the dependent variable. Only the data for females are shown in this figure.

In the simultaneous models (Figure 6–9 and Additional file 8-11), all 52 lifestyle factors were included as covariates, allowing the detection of independent variables associated with DNAmAge, Dunedin PACE, and AgeAccel. In males, fruit intake, late-night snacking, direct sunlight exposure, and water intake were positively associated with specific clocks. In contrast, vegetable intake showed a clear negative association with GrimAge-related clocks, whereas sauna use and stone spa bathing exhibited a strong association with multiple clocks. When compared with the single-factor models, simultaneous models revealed that the associations of processed food intake, artificial sweetener use, and smoking-related behaviors with epigenetic clocks were diminished, whereas late-night snacking, fruit intake, sunlight exposure, and thermal relaxation were still associated with epigenetic aging in males. In females, HIIT maintained a strong association with multiple clocks, including the Horvath Clock and GrimAge v2 after adjusting for all other lifestyle factors. Sleep quality and stretching also retained negative associations after adjustment. Conversely, electronic cigarette use, caloric restriction frequency, nut intake, water intake, and coffee consumption were positively associated with AgeAccel in specific clocks, such as Horvath, GrimAge v2, and Hannum. Compared with the models with single lifestyle factors, smoking-related exposure showed reduced effect sizes, whereas caloric restriction, nut and water intake, coffee consumption, and high-intensity exercise had strong associations.

**Figure 6.**
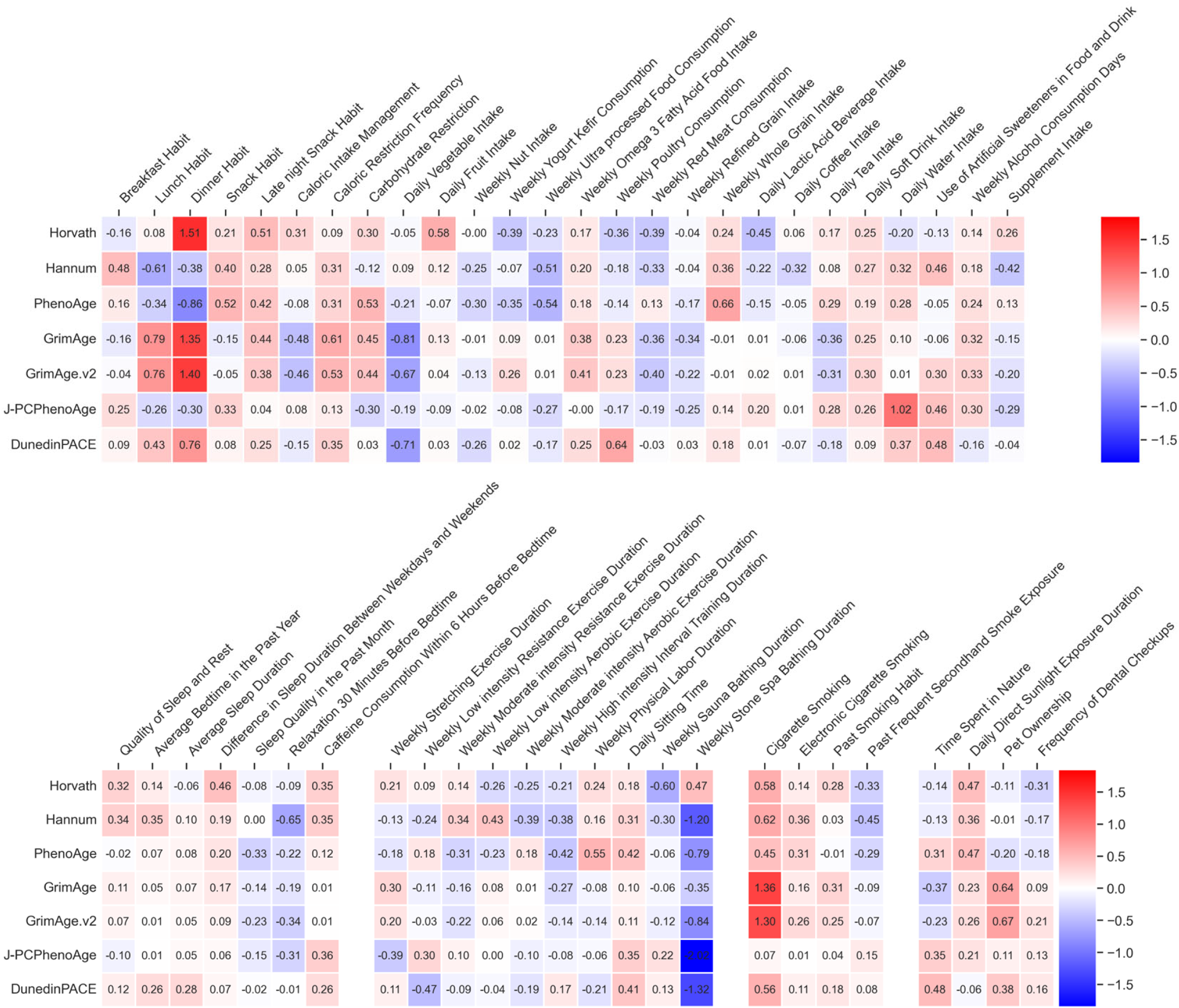
Heatmap of effect sizes for associations between seven DNAmAge indicators and all 52 lifestyle-related factors in males. Each cell represents the standardized effect size derived from multivariable linear regression models, in which all lifestyle factors were entered as explanatory variables and a DNAmAge indicator (Horvath, Hannum, PhenoAge, GrimAge, GrimAge v2, J-PCPhenoAge, or DunedinPACE) was used as the dependent variable. Only the data for males are shown in this figure.

**Figure 7.**
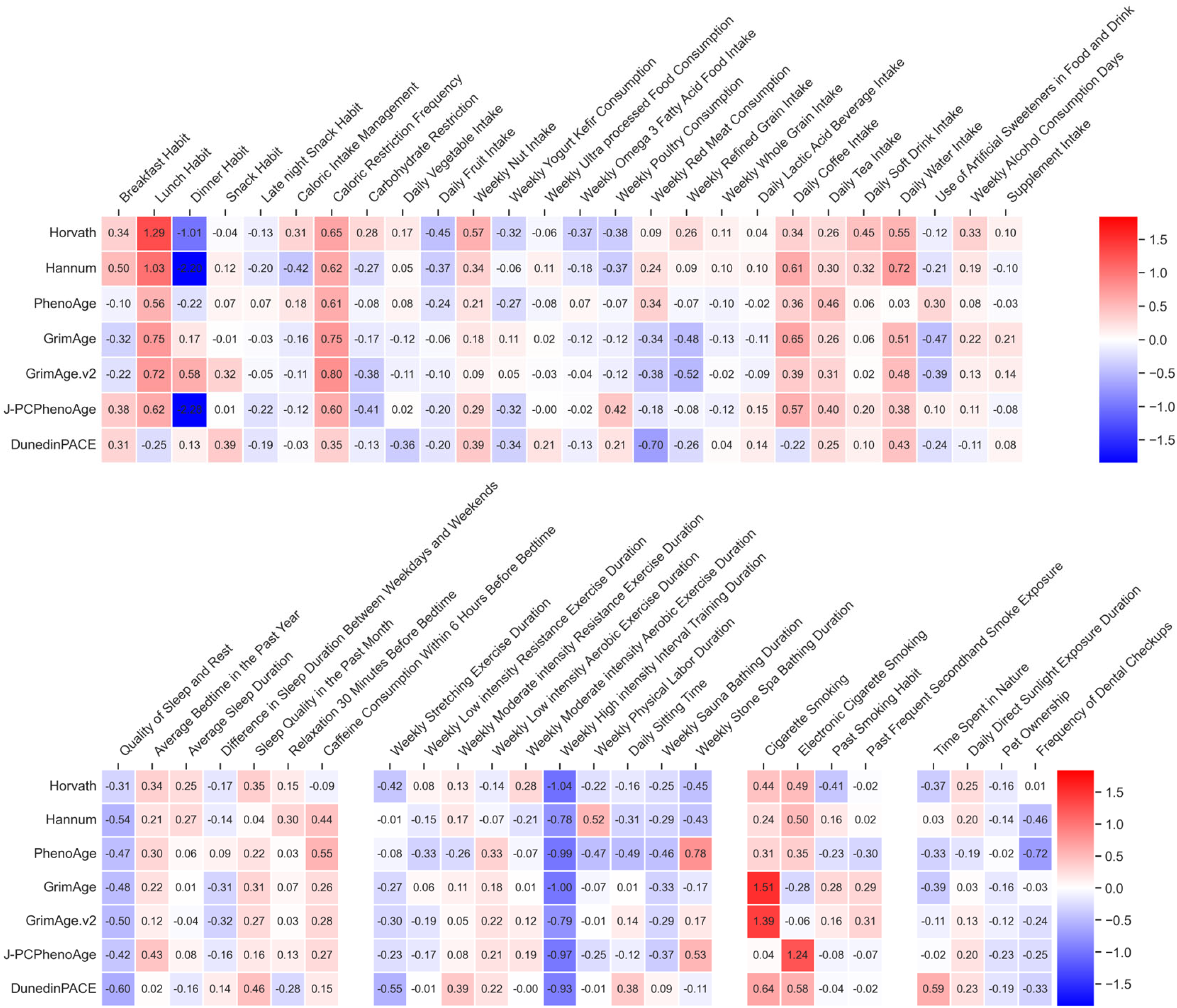
Heatmap of effect sizes for associations between seven DNAmAge indicators and all 52 lifestyle-related factors in females. Each cell represents the standardized effect size derived from multivariable linear regression models, in which all lifestyle factors were entered as explanatory variables and a DNAmAge indicator (Horvath, Hannum, PhenoAge, GrimAge, GrimAge v2, J-PCPhenoAge, or DunedinPACE) was used as the dependent variable. Only the data for females are shown in this figure.

**Figure 8.**
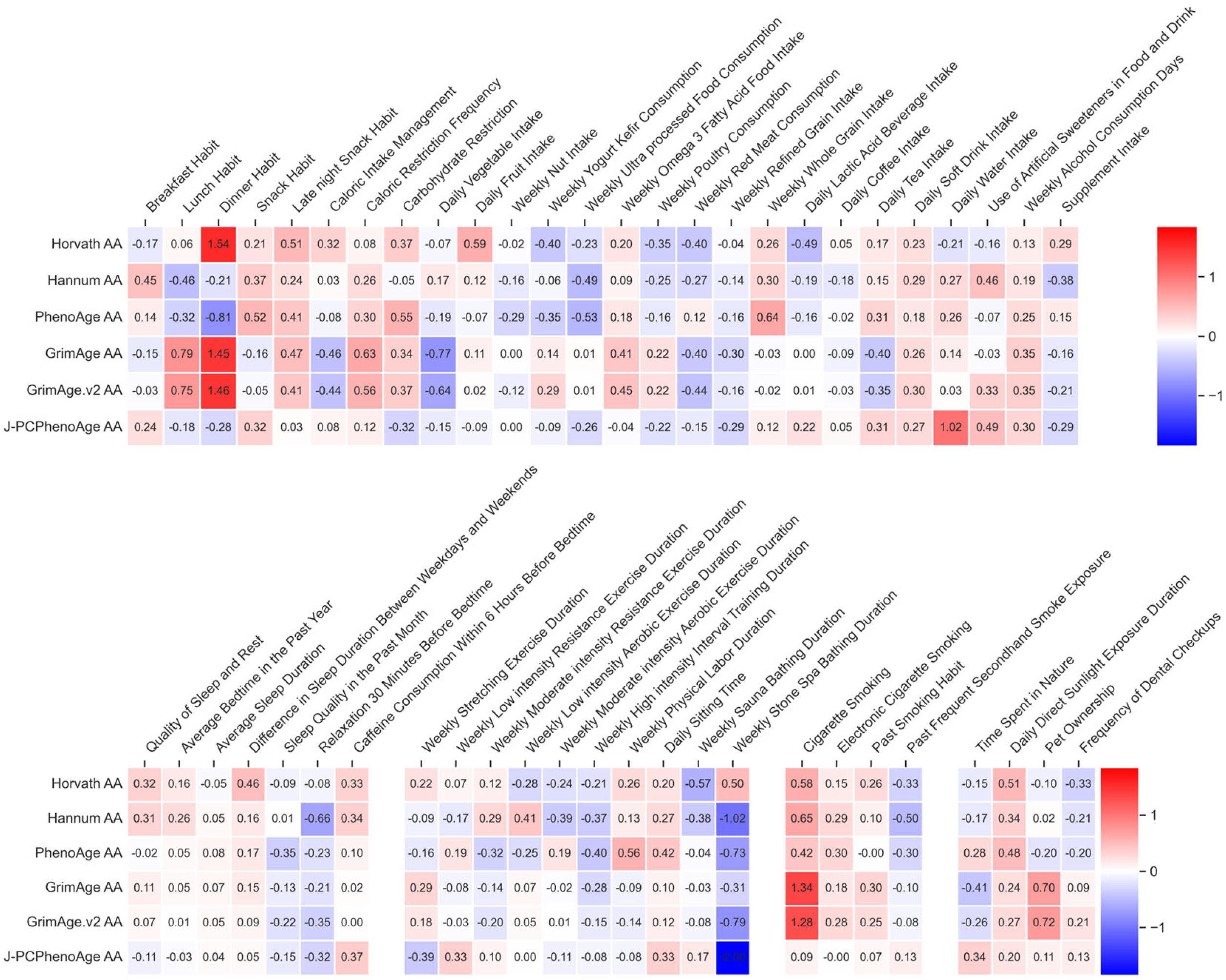
Heatmap of effect sizes for associations between six AgeAccel indicators and all 52 lifestyle-related factors in males. Each cell represents the standardized effect size derived from multivariable linear regression models in which all lifestyle factors were entered as explanatory variables and an AgeAccel indicator (Horvath AA, Hannum AA, PhenoAge AA, GrimAge AA, GrimAge v2 AA, or J-PCPhenoAge AA) was used as the dependent variable. Only the data for males are shown in this figure.

**Figure 9.**
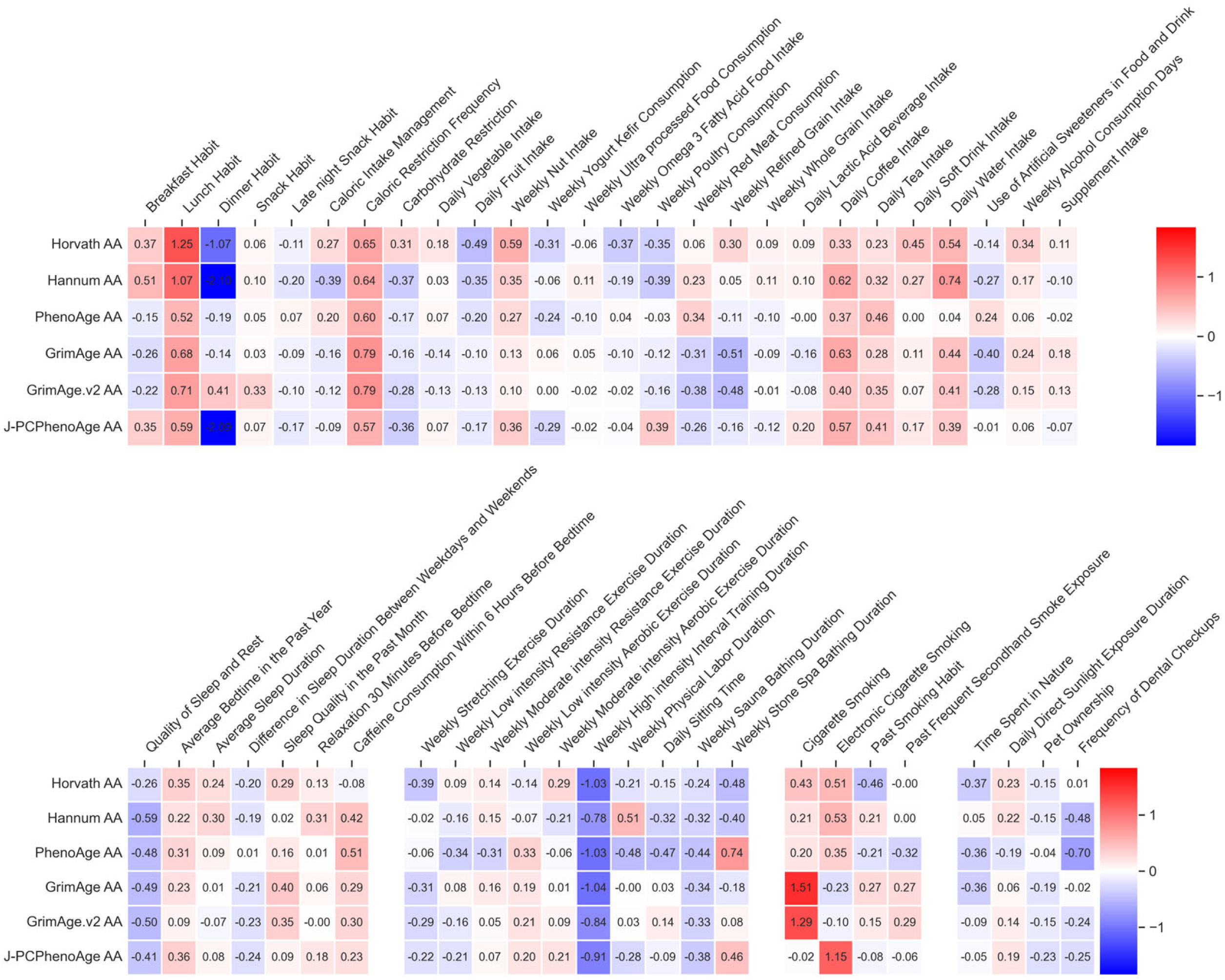
Heatmap of effect sizes for associations between six AgeAccel indicators and all 52 lifestyle-related factors in females. Each cell represents the standardized effect size derived from multivariable linear regression models in which all lifestyle factors were entered as explanatory variables and an AgeAccel indicator (Horvath AA, Hannum AA, PhenoAge AA, GrimAge AA, GrimAge v2 AA, or J-PCPhenoAge AA) was used as the dependent variable. Only the data for females are shown in this figure.

## Discussion

In this study, we examined the associations between 52 lifestyle factors and 13 epigenetic aging indicators (six DNAmAge measures, Dunedin PACE, and six AgeAccel measures) in males and females separately. We found that multiple lifestyle domains, including dietary behaviors, smoking-related exposures, beverage consumption, exercise habits, and thermal relaxation practices, exhibited strong accelerating or decelerating effects on at least one epigenetic aging indicator. As shown in Figure 10 and Additional file 12, these lifestyle factors demonstrated consistent effect size patterns across both single-factor and simultaneous-entry models.

**Figure 10.**
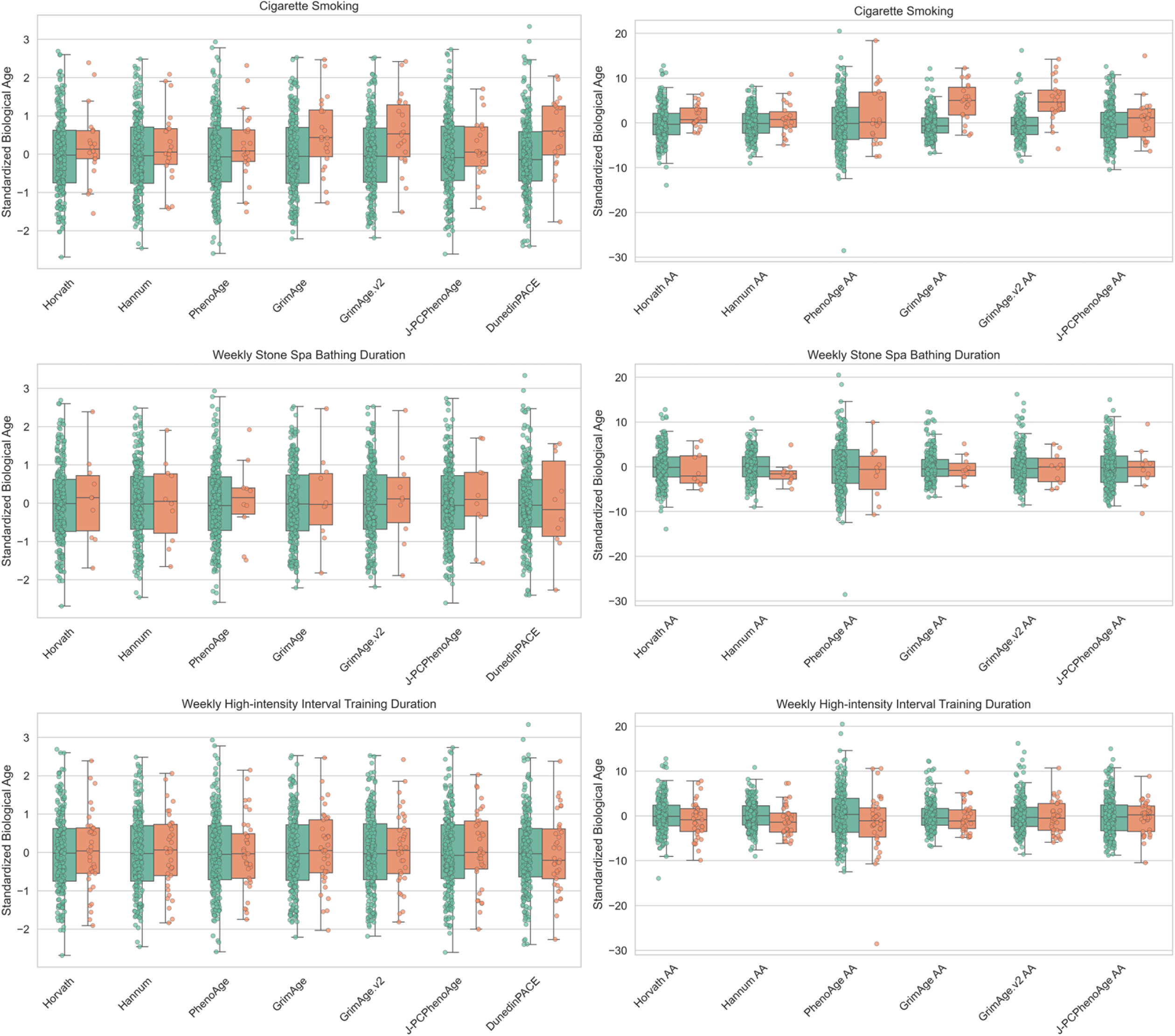
Comparison of DNAmAge and AgeAccel values according to key lifestyle factors. Boxplots illustrate the distributions of seven DNAmAge measures (left panels) and six AgeAccel measures (right panels), stratified by the presence (orange) or absence (green) of three lifestyle factors: cigarette smoking, weekly stone spa bathing duration, and weekly HIIT duration.

There were clear sex differences in the overall structure of the influential lifestyle factors. In males, smoking behaviors were positively associated with both DNAmAge and AgeAccel, whereas thermal relaxation behaviors such as sauna use and stone spa bathing showed a negative association. In females, electronic cigarette use, caloric and carbohydrate restrictions, nut intake, and water and coffee consumption were positively associated with multiple epigenetic clocks, whereas HIIT consistently showed negative associations. These patterns indicate that lifestyle factors influence epigenetic aging in a sex-specific manner, highlighting the need for sex-specific intervention strategies in personalized healthcare.

When the results were organized by the six DNAmAges, the Dunedin PACE, and the six AgeAccel indicators, each metric exhibited a distinct pattern of association with lifestyle behaviors. The Horvath Clock showed the broadest range of associations, including both positive and negative associations with dietary behaviors and smoking. The Hannum Clock generally demonstrated smaller effect sizes but displayed a selective association with beverage consumption, particularly coffee intake, in females. PhenoAge consistently indicated lower aging estimates among individuals who engaged in high-intensity exercise, especially in females. GrimAge was strongly associated with smoking-related behaviors and dietary practices, such as late-night eating and fruit intake. GrimAge v2 demonstrated the greatest variability in effect sizes and showed marked associations with electronic cigarette use and thermal relaxation behaviors. J-PCPhenoAge exhibited associations with only a limited set of behaviors. The most notable behaviors were water intake, in simultaneous-entry models, highlighting a pattern that differs from other clocks. The Dunedin PACE generally showed small effect sizes and limited associations with lifestyle factors, although weak associations with smoking and physical activity were observed.

Our findings regarding the impact of smoking and physical activity on the six DNAmAges, Dunedin PACE, and six AgeAccel indicators are largely consistent with previous large-scale Japanese cohort studies, such as the J-MICC study [27], which reported negative associations between physical activity and epigenetic aging. However, the present study extended these findings by examining a broader array of 52 lifestyle factors in a clinical population, revealing novel associations between sauna use, stone spa bathing, and specific dietary behaviors with each epigenetic indicator. Furthermore, while previous studies relied primarily on clocks developed using data derived from Western populations, our inclusion of population-specific J-PCPhenoAge data provides a complementary perspective, suggesting that suitable clocks for the target population may refine the detection of lifestyle-related aging signals.

Some lifestyle factors with highly imbalanced response distributions, such as lunch and dinner habits, require cautious interpretation as such skewness may limit the reliability and stability of the regression estimates. Correlation analyses further clarified the biological structure of the clocks: First- and second-generation DNAmAge indicators were strongly intercorrelated, whereas Dunedin PACE, reflecting aging pace, showed weaker associations, consistent with prior evidence that aging level and pace represent different biological constructs [28]. AgeAccel indicators displayed weaker overall correlations, supporting the interpretation that residualized measures capture more heterogeneous aging processes. The Japanese-specific J-PCPhenoAge showed distinct correlation patterns, suggesting a population-specific methylation signature.

As shown in Table 4, while the Horvath clock overestimated the biological age of the Japanese participants, with approximately 89% classified as older than their chronological age, J-PCPhenoAge showed a more balanced distribution. This discrepancy and the distinct J-PCPhenoAge pattern may reflect the successful removal of population-specific bias through learning in the Japanese population. Furthermore, an independent study conducted in a Korean cohort demonstrated that J-PCPhenoAge outperformed epigenetic clocks that were developed primarily in Western populations [29]. Specifically, J-PCPhenoAge showed stronger correlations with chronological age than the PCHorvath, PCHannum, and PCPhenoAge and showed slightly stronger associations with health outcomes such as Type II diabetes and hypertension. These findings in an independent cohort provide additional support for the validity of J-PCPhenoAge.

These findings have important implications for personalized healthcare and public health. The selective associations of the clocks suggest that multiple indicators should be used jointly rather than relying on a single metric. Lifestyle domains with consistent effects such as smoking-related exposure, meal timing, and high-intensity exercise are promising targets for precision lifestyle interventions. Moreover, clear sex-related differences emphasize the relevance of personalized strategies in behavioral change programs. In the Japanese society, where healthy life expectancy and rising medical costs remain major concerns, epigenetic age may serve as an intuitive metric for communicating health risks and supporting behavioral modification. The distinctive behavioral clock associations observed for J-PCPhenoAge highlight the importance of developing population-specific biological age measures.

Some limitations should be noted. The observed associations can not necessarily be interpreted as causal relationships, and the participants may differ from the general Japanese population in terms of health awareness, which potentially limits generalizability. Lifestyle factors were self-reported and dichotomized, which may obscure dose–response relationships. Unmeasured confounding factors remain a challenge. Future research should incorporate longitudinal analyses, controlled interventions, and cross-cultural comparisons to refine our understanding of lifestyle determinants of epigenetic aging. In summary, our findings provide comprehensive evidence linking diverse lifestyle behaviors to multiple dimensions of biological aging in Japanese adults, thereby offering a foundation for personalized healthcare and the development of biologically informed public health strategies.

## Conclusions

Our findings reveal multifaceted associations between lifestyle behaviors and epigenetic aging and provides foundational evidence for advancing personalized healthcare based on biological age. Future research will require a longitudinal design, replication in multicultural populations, and intervention studies to evaluate the modifiability of epigenetic aging through lifestyle improvements.

## Data Availability

A full list of the lifestyle-related questionnaire items is provided in Additional file 1. The response to all lifestyle questionnaire items listed in Additional file 1 is summarized in Additional file 2 and 3. Estimated regression coefficients, 95% confidence intervels, and p-values are provided in Additional file 4-11. All other data supporting the findings of this study are not publicly available due to ethical and privacy considerations but are available from the corresponding author upon reasonable request.

## List of abbreviations

DNAm: DNA methylation;
DNAmAge: DNA methylation age;
J-PCPhenoAge: PC-PhenoAge for Japanese Population;
HIIT: high-intensity interval training;
SD: standard deviation

## Declarations

### Ethics approval and consent to participate

This study was conducted in accordance with the Declaration of Helsinki. The study protocol was reviewed and approved by the Ethics Committee of the Asia–Oceania Association Anti-Aging Promotion (approval number: AOAAPA22-001). All participants were informed of the study objectives and procedures, and written informed consent was obtained from all individuals prior to participation.

### Consent for publication

Not applicable.

### Authors’ contributions

Conceptualization: RN, SH, and HY;

Study design and data collection: RN, SH, and HY;

DNA methylation analysis: GY and TS;

Statistical analysis: GY and TS;

Interpretation of results: TS, RN, SH, and HY;

Manuscript drafting: TS;

Manuscript review and editing: all authors.

All authors have read and approved the final manuscript.

## Acknowledgements

The authors thank Yui Tomo for advice regarding the statistical methods. The authors thank Yasutaka Hayashida, M.D. (SAWAKO CLINIC × YS) for the collection of the Japanese blood samples that formed the foundation of this study. The authors thank Editage (www.editage.com) for providing English language editing assistance. The contributions of these individuals were indispensable for the successful execution of this study.

## Additional files

### File name: Additional file 1

File format: xlsx

Title of data: **Questionnaire Items for Lifestyle-Related Factors.**

Description of data: This table provides a full list of the lifestyle-related questionnaire items. The items were categorized into six domains—Food, Sleep, Exercise, Smoking, Others, and Covariate-related items—each of which was designed to capture behavioral factors previously reported to influence metabolic health, chronic disease risk, and epigenetic aging. The table includes the original wording of each question, categorical response options, and the conceptual lifestyle domain to which each item belongs. These items served as the basis for the subsequent descriptive analyses and variable coding.

### File name: Additional file 2

File format: xlsx

Title of data: **Distribution of Raw Questionnaire Responses.**

Description of data: This table summarizes the raw responses to all lifestyle questionnaire items listed in Additional file 1. These raw responses were subsequently processed into binary variables for regression modeling.

### File name: Additional file 3

File format: xlsx

Title of data: **Binary Coding of Lifestyle-Related Questionnaire Responses.**

Description of data: This table presents the dichotomized versions of all 52 lifestyle-related questionnaire items. Each item from the raw questionnaire responses was converted into a binary variable (0 or 1) for use as an explanatory variable in the regression models assessing associations with the DNAmAge and AgeAccel indicators.

### File name: Additional file 4

File format: xlsx

Title of data: **Estimated regression coefficients, 95% confidence intervals, and p-values in the single-factor model with DNAmAge as the dependent variable (Male).**

Description of data

### File name: Additional file 5

File format: xlsx

Title of data: **Estimated regression coefficients, 95% confidence intervals, and p-values in the single-factor model with DNAmAge as the dependent variable (Female).**

Description of data

### File name: Additional file 6

File format: xlsx

Title of data: **Estimated regression coefficients, 95% confidence intervals, and p-values in the single-factor model with AgeAccel as the dependent variable (Male).**

Description of data

### File name: Additional file 7

File format: xlsx

Title of data: **Estimated regression coefficients, 95% confidence intervals, and p-values in the single-factor model with AgeAccel as the dependent variable (Female).**

Description of data

### File name: Additional file 8

File format: xlsx

Title of data: **Estimated regression coefficients, 95% confidence intervals, and p-values in the simultaneous model with DNAmAge as the dependent variable (Male).**

Description of data

### File name: Additional file 9

File format: xlsx

Title of data: **Estimated regression coefficients, 95% confidence intervals, and p-values in the simultaneous model with DNAmAge as the dependent variable (Female).**

Description of data

### File name: Additional file 10

File format: xlsx

Description of data

**Title of data: Estimated regression coefficients, 95% confidence intervals, and p-values in the simultaneous model with AgeAccel as the dependent variable (Male).**

### File name: Additional file 11

File format: xlsx

Title of data: **Estimated regression coefficients, 95% confidence intervals, and p-values in the simultaneous model with AgeAccel as the dependent variable (Female).**

Description of data:

### File name: Additional file 12

**File format:** tif

**Title of data: Comparison of DNAmAge and AgeAccel values according to other lifestyle factors**

**Description of data:** Boxplots illustrate the distribution of seven DNAmAge measures (left panels) and six AgeAccel measures (right panels), stratified by the presence (orange) or absence (green) of three lifestyle factors: caloric restriction frequency, daily coffee intake, daily water intake, electronic cigarette smoking, late-night snack habits, and weekly nut intake.

