## Supplementary figures and images for "Comprehensive Evaluation of Associations between Lifestyle Factors and Multiple Epigenetic Aging Indicators in the Japanese Population: A cross-sectional study"

### Additional file 12

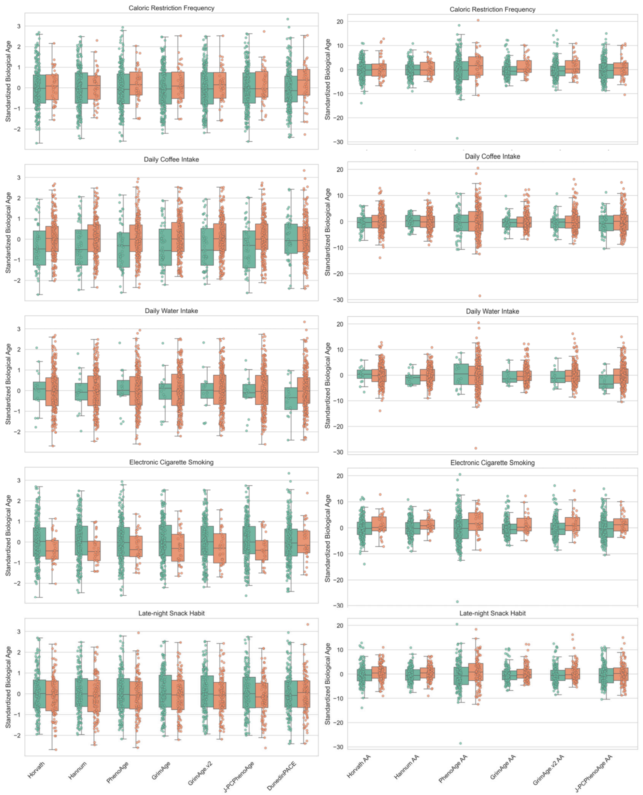

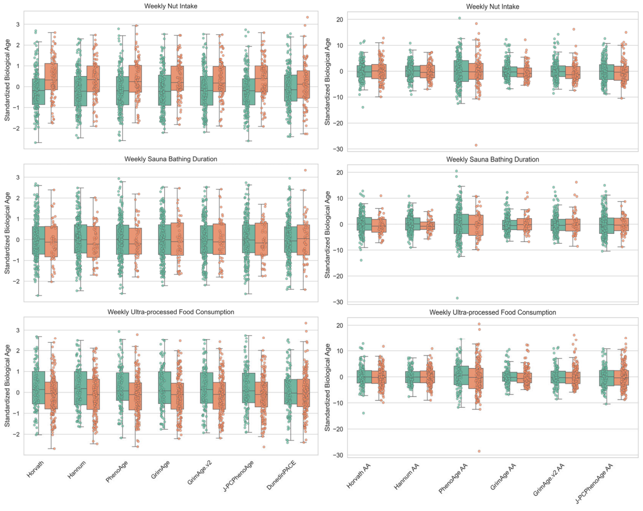
